# COVID-19 clinical characteristics, and sex-specific risk of mortality: Systematic Review and Meta-analysis

**DOI:** 10.1101/2020.03.24.20042903

**Authors:** Mohammad Javad Nasiri, Sara Haddadi, Azin Tahvildari, Yeganeh Farsi, Mahta Arbabi, Saba Hasanzadeh, Parnian Jamshidi, Mukunthan Murthi, Mehdi S. Mirsaeidi

## Abstract

**Objectives:** The rapidly evolving coronavirus disease 2019 (COVID-19), was declared a pandemic by the World Health Organization on March 11, 2020. It was first detected in the city of Wuhan in China and has spread globally resulting in substantial health and economic crisis in many countries. Observational studies have partially identified the different aspects of this disease. Up to this date, no comprehensive systematic review for the clinical, laboratory, epidemiologic and mortality findings has been published. We conducted this systematic review and meta-analysis for a better understanding of COVID-19.

**Methods:** We reviewed the scientific literature published from January 1, 2019 to March 3, 2020. Statistical analyses were performed with STATA (version 14, IC; Stata Corporation, College Station, TX, USA). The pooled frequency with 95% confidence intervals (CI) was assessed using random effect model. Publication bias was assessed and p <0.05 was considered a statistically significant publication bias.

**Results:** Out of 1102 studies, 32 satisfied the inclusion criteria. A total of 4789 patients with a mean age of 49 years were evaluated. Fever (83.0%, CI 77.5 to 87.6), cough (65.2%, CI 58.6 to 71.2) and myalgia/fatigue (34.7, CI 26.0 to 44.4) were the most common symptoms. The most prevalent comorbidities were hypertension (18.5 %, CI 12.7 to 24.4) and Cardiovascular disease (14.9 %, CI 6.0 to 23.8). Among the laboratory abnormalities, elevated C-Reactive Protein (CRP) (72.0% (CI 54.3 to 84.6) and lymphopenia (50.1%, CI 38.0 to 62.4) were the most common findings. Bilateral ground-glass opacities (66.0%, CI 51.1 to 78.0) was the most common CT-Scan presentation. Pooled mortality rate was 6.6%, with males having significantly higher mortality compared to females (OR 3.4; 95% CI 1.2 to 9.1, P = 0.01).

**Conclusion:** COVID-19 commonly presented with a progressive course of cough and fever with more than half of hospitalized patients showing leukopenia or a high CRP on their laboratory findings. Mortality associated with COVID19 was higher than that reported in studies in China with Males having a 3-fold higher risk of mortality in COVID19 compared to females.

**Summary box:** *What is already known in this topic:*

- COVID-19 was declared a pandemic by the World Health Organization on March 11, 2020.
- Many observational studies have separately dealt with different clinical and epidemiologic features of this new and rapidly evolving disease.
- Very few systematic reviews about COVID-19 have been done and there was still a need for a systematic review and meta-analysis related to the clinical findings and the mortality of the disease in order to have a better understanding of COVID-19.
- Previous reports have indicated that older age and presence of multiple comorbidities are associated with increased mortality.

*What this study adds:*

- The mortality rate in our study for hospitalized COVID-19 patients was 6.6% and males had around 3-fold higher risk of mortality compared to females (OR 3.4; 95% CI 1.2-9.1, P = 0.01).
- These findings could indicate the need for more aggressive treatment of COVID-19 in males.

## Introduction

Facing an immediate crisis by the novel coronavirus, Severe Acute Respiratory Syndrome Coronavirus 2, (SARS-CoV-2), which has been called the once in a century pathogen requires a global response to this outbreak(1). The disease caused by this virus has been named “coronavirus disease 2019” (COVID-19) by the World Health Organization. As of now, more than 168 countries have reported COVID19 patients. Given the increasing number of countries infected with SARS-CoV-2, WHO declared COVID19 a pandemic on March 11, 2020.(2) The SARS-CoV-2 virus is a betacoronavirus and like the Middle East Respiratory Syndrome virus (MERS-CoV) and SARS-CoV that caused the previous respiratory syndrome outbreaks, belongs to the family of coronaviruses. However, this is the first pandemic caused by a member of the coronavirus family (3).

COVID19 started in China in December 2019 when a cluster of patients with pneumonia of unknown cause was identified in the city of Wuhan. Since then, it has infected hundreds of thousands of people around the world and resulted in more than 13000 deaths up to this date (4). Despite governmental travel restrictions in many countries, the confirmed number of new cases has been rising globally. The international community has asked for at least US$675 million for preparedness and protection of states with weaker health systems (5).

In the previous two outbreaks of coronaviral respiratory illness, namely Severe Acute Respiratory Illness (SARS) and Middle East Respiratory Illness (MERS), gender-based difference in mortality was observed. In SARS, younger males were at twice the risk of death compared to females and the difference in mortality reduced with older age(6). The case fatality rate observed in males was twice that of females in MERS (7). The effect of sex on COVID-19 mortality was unknown. We evaluated this risk for COVID-19 patients as well.

The novelty of COVID19 has raised many questions about the epidemiology of the disease, clinical and laboratory methods of diagnosis as well as therapeutic measures. Thus far, many observational studies have been dealing with these features separately, however, there is still a necessity for more systematic reviews specially to understand role of the sex in mortality of COVID19. In this meta-analysis study, we reviewed the published literature from January 1, 2019 to March 3, 2020 to provide an overview for a better understanding of COVID-19.

## Methods

### Search strategy

We searched Pubmed/Medline, Embase, Web of Science and the Cochrane Library for studies published from January 1, 2019 to March 3, 2020. The search strategy was based on the following key-words: COVID-19, severe acute respiratory syndrome coronavirus 2, novel coronavirus, SARS-CoV-2, nCoV disease, SARS2, COVID19, Wuhan coronavirus, Wuhan seafood market pneumonia virus, 2019-nCoV, coronavirus disease-19, coronavirus disease 2019, 2019 novel coronavirus and Wuhan pneumonia. Lists of references of selected articles and relevant review articles were hand-searched to identify further studies. This study was conducted and reported according to the PRISMA guidelines (8). The study did not require any ethics committee approval. This research was done without patient involvement. Patients were not invited to comment on the study design and were not consulted to develop patient relevant outcomes or interpret the results. Patients were not invited to contribute to the writing or editing of this document for readability or accuracy.

### Study Selection

The records found through database searching were merged and the duplicates were removed using EndNote X7 (Thomson Reuters, New York, NY, USA). Two reviewers (YF and PJ) independently screened the records by title and abstract to exclude those not related to the current study. The full texts of potentially eligible records were retrieved and evaluated by a third reviewer (AT). Included studies met the following inclusion criteria: (i) patients were confirmed and diagnosed with RT-PCR as suggested by WHO; (ii) The raw data for clinical, radiological and laboratory findings were included; and (iii) the outcomes were addressed. Studies with insufficient information about patients’ characteristics and outcomes were excluded. Case reports, reviews, and animal studies were also excluded. Only studies written in English were selected.

### Data extraction and quality assessment

A data extraction form was designed by two reviewers (AZ and SH). These reviewers extracted the data from all eligible studies and differences were resolved by consensus. The following data were extracted: first author name; year of publication; type of study, country/ies where the study was conducted; distribution of age and sex in the population, number of patients investigated, data for clinical, radiological and laboratory findings, and outcomes.

### Data Synthesis and Analysis

Statistical analyses were performed with STATA (version 14, IC; Stata Corporation, College Station, TX, USA). The pooled frequency with 95% confidence intervals (CI) was assessed using random effect model. The between-study heterogeneity was assessed by Cochran’s Q and the I2 statistic. Publication bias was assessed statistically by using Begg’s and Egger’s tests (p<0.05 was considered indicative of statistically significant publication bias).

### Quality assessment

The checklist provided by the Joanna Briggs Institute (JBI) was used to perform quality assessment(9).

## Results

The search yielded 1102 publications, of which 259 potentially eligible studies were identified for full-text review, resulting in 32 studies fulfilling the inclusion criteria (Figure 1) (Table1). The mean age of the patients was 49.0 years and 4789 patients were included. Based on JBI tool, the included studies had a low risk of bias.

**Table 1.** Characteristics of the included studies.

| **First Author** | **Country** | **Published time** | **Type of study** | **Mean age** | **Male/Female** | **Nationality** | **No. of patients** | **Diagnostic methods** |
| --- | --- | --- | --- | --- | --- | --- | --- | --- |
| S. Hui(32) | China | 14, Jan, 2020 | Case series | NR | NR | Chinese | 41 | RT-PCR/CT-scan |
| Xia(33) | China | 26, Feb, 2020 | Case series | 54.5 | 21M, 9F | Chinese | 30 | RT-PCR |
| Wei Xu(34) | China | 13, Feb, 2020 | Case series | 41 | M35, F27 | Chinese | 62 | RT-PCR |
| Wei Zhang(35) | China | 7, Feb, 2020 | Case series | NR | NR | Chinese | 178 | RT-PCR |
| Kai-Wang To(36) | China | 12, Feb, 2020 | Case series | 62.5 | 7M, 5F | Chinese | 12 | RT-PCR |
| Zou(37) | China | 19, Feb, 2020 | Correspondence | 59 | 9M ,9F | Chinese | 18 | RT-PCR |
| Hoehl(38) | Germany | 3, Mar, 2020 | Correspondence | 35 | NR | German | 126 | RT-PCR/CT-scan |
| Yang Pan(39) | China | 24, Feb, 2020 | Correspondence | NR | NR | Chinese | 82 | RT-PCR/CT-scan |
| Tang(40) | China | 19, Feb, 2020 | Cross-sectional | 54 | 98M, 85F | Chinese | 183 | RT-PCR |
| Chung(41) | China | 4, Feb, 2020 | Cross-sectional | 51 | M13, F8 | Chinese | 21 | RT-PCR/CT-scan |
| Yicheng Fang2(42) | China | 19, Feb, 2020 | Cross-sectional | 45 | 29M, 22F | Chinese | 51 | RT-PCR/CT-scan |
| Guan(43) | China | 28, Feb, 2020 | Cross-sectional | 47 | 640M,459F | Chinese | 1099 | RT-PCR/CT-scan |
| Huang(44) | China | 24, Jan, 2020 | Cross-sectional | 49 | 30M,11F | Chinese | 41 | RT-PCR |
| Kui Liu(45) | China | 7, Feb, 2020 | Cross-sectional | 57 | 61M,76F | Chinese | 137 | RT-PCR |
| Qun Li(46) | China | 29, Jan, 2020 | Cross-sectional | 52 | M238, F187 | Chinese | 425 | RT-PCR/CT-scan |
| Yingxia Liu(47) | China | 9, Feb, 2020 | Cross-sectional | 53.6 | 8M, 4F | Chinese | 12 | RT-PCR/CT-scan |
| Dawei Wang(48) | China | 7, Feb, 2020 | Cross-sectional | 56 | 75M , 63F | Chinese | 138 | RT-PCR/CT-scan |
| Jian Wu(49) | China | 29, Feb, 2020 | Cross-sectional | 46 | 39M, 41F | Chinese | 80 | RT-PCR |
| Jin-jin Zhang(10) | China | 19, Feb, 2020 | Cross-sectional | 57 | 71M,69F | Chinese | 140 | RT-PCR |
| Ai(50) | China | 26, Feb, 2020 | Cross-sectional | 48.5 | M467, F547 | Chinese | 1014 | RT-PCR/CT scan |
| Feng Pan(51) | China | 13, Feb, 2020 | Cross-sectional | 40 | 6M, 15F | Chinese | 21 | RT-PCR/CT-scan |
| Heshui Shi2(52) | China | 24, Feb, 2020 | Cross-sectional | 49.5 | 42M, 39F | Chinese | 81 | RT-PCR/CT-scan |
| Yang(53) | China | 21, Feb, 2020 | Cross-sectional | 59.7 | 35M, 17F | Chinese | 52 | RT-PCR |
| Bajema(54 ) | China | 4, Feb, 2020 | Cross-sectional | NR | 115M, 95F | Chinese | 210 | RT-PCR/CT-scan |
| Bernheim(55) | China | 20, Feb, 2020 | Cross-sectional | 45.3 | 61M, 60F | Chinese | 121 | RT-PCR |
| Nanshan Chen(56) | China | 15, Feb, 2020 | Cross-sectional | 55.5 | 67M, 32F | Chinese | 99 | RT-PCR |
| Yueying Pan(57) | China | 13, Feb, 2020 | Cross-sectional | 45 | 33M, 30F | Chinese | 63 | RT-PCR |
| Yu-Huan Xu(58) | China | 21, Feb, 2020 | Cross-sectional | 44 | 29M, 21F | Chinese | 50 | RT-PCR/CT-scan |
| Xi Xu(59) | China | 28, Feb, 2020 | Cross-sectional | 50 | 39M, 51F | Chinese | 90 | RT-PCR |
| De Chang(60) | China | 7, Feb, 2020 | Research letter | 34 | 10M, 3F | Chinese | 13 | RT-PCR/CT-scan |
| Weilie Chen(61) | China | 26, Feb, 2020 | Research letter | NR | NR | Chinese | 85 | RT-PCR/CT-scan |
| Kwok(62) | China | 7, Feb, 2020 | Research letter | 59.8 | 9M, 5F | Chinese | 14 | RT-PCR/CT-scan |

**Figure 1.**
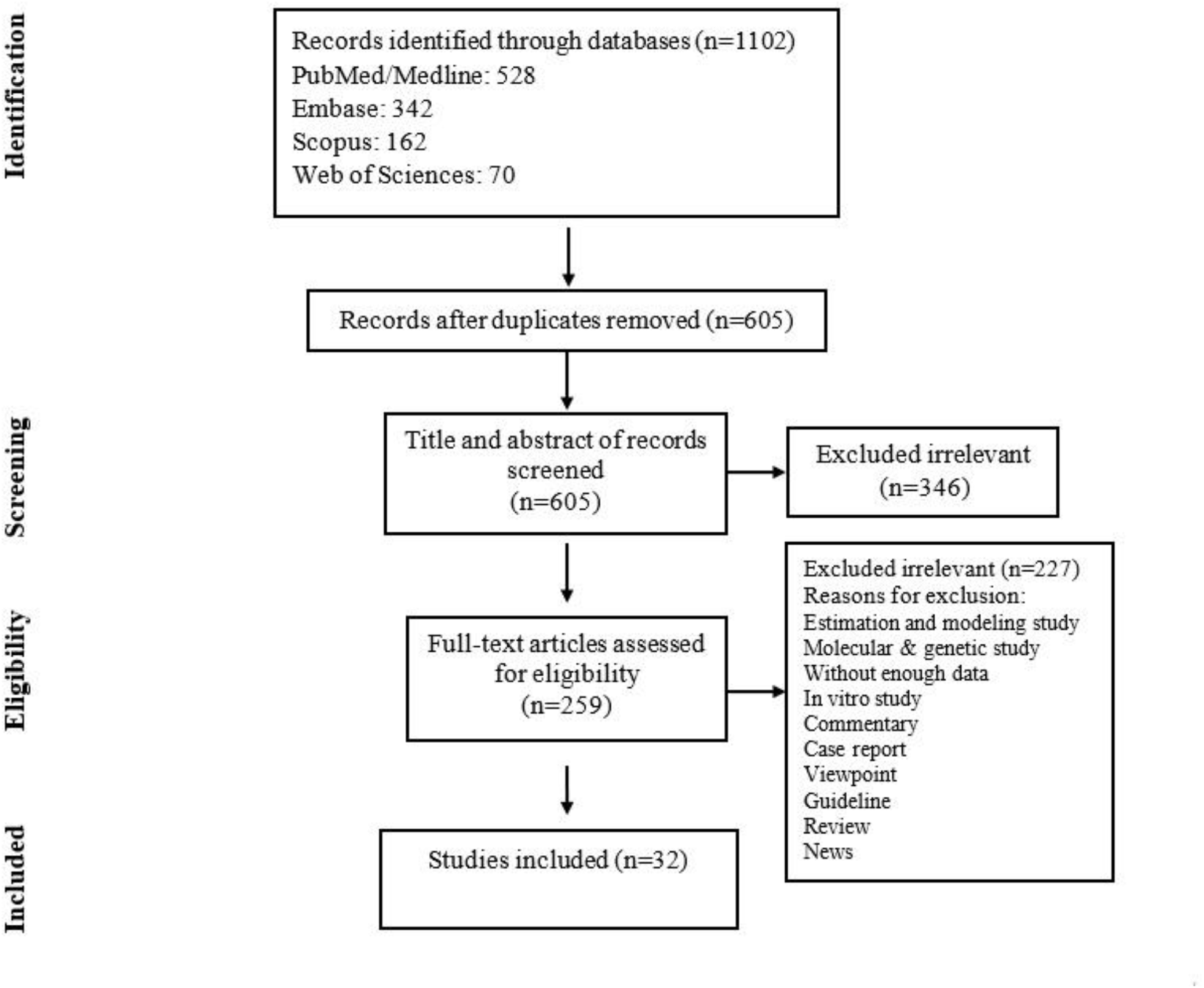
Flow chart of study selection for inclusion in the systematic review and meta-analysis.

### Clinical manifestations

The most common signs and symptoms were fever (83.0%, CI 77.5-87.6), cough (65.2%, CI 58.6-71.2), dyspnea (27.4%, CI 19.6-35.2), myalgia/fatigue (34.7, CI 26.0-44.4) followed by hemoptysis (2.4%, CI 0.8-6.7), diarrhea (5.7%, CI 3.8-8.6) and nausea/vomiting (5.0 %, CI 2.3-10.7). Sputum production (17.2%, CI 10.8-26.4) was a relatively common symptom. (Table 2).

**Table 2.** Meta-analysis of comorbidities.

|  | Number of studies | Pooled frequency (95% CI) | n/N* | Publication bias (*p* value) | Heterogeneity test |  |
| --- | --- | --- | --- | --- | --- | --- |
|  |  |  |  |  | I<sup>2</sup> (%) | *p* value |
| Smoking | 4 | 8.0 (2.3-13.6) | 172/1332 | 0.06 | 100 | 0.00 |
| Hypertension | 9 | 18.5 (12.7-24.4) | 306/1800 | 0.98 | 100 | 0.00 |
| Cardiovascular disease | 12 | 14.9 (6.0-23.8) | 178/2031 | 0.72 | 100 | 0.00 |
| Diabetes | 11 | 10.8 (8.3-13.3) | 166/1932 | 0.39 | 100 | 0.00 |
| Pulmonary disease | 12 | 3.4 (0.8-6.0) | 39/2031 | 0.72 | 100 | 0.00 |
| Malignancies | 9 | 2.8 (0.8-4.8) | 33/1816 | 0.74 | 100 | 0.00 |
| Chronic liver disease | 7 | 8.1 (4.6-11.6) | 29/546 | 0.45 | 100 | 0.00 |
| Renal disease | 6 | 4.4 (0.24-8.6) | 17/1472 | 0.33 | 100 | 0.00 |

\*n, number of patients with comorbidity; N, total number of patients.

### Comorbidities

The most common comorbidities were respectively hypertension (18.5 %, CI 12.7-24.4), cardiovascular diseases (14.9 %, CI 6.0-23.8), diabetes (10.8 %, CI 8.3-13.3), chronic liver disease (8.1, CI 4.6-11.6) and smoking (8.0%, CI 2.3-13.6) (Table 3).

**Table 3.** Meta-analysis of clinical manifestations.

|  | Number of studies | Pooled frequency (95% CI) | n/N* | Publication bias (p value) | Heterogeneity test |  |
| --- | --- | --- | --- | --- | --- | --- |
|  |  |  |  |  | I<sup>2</sup> (%) | p value |
| Fever | 21 | 83.0 (77.5-87.6) | 2073/2465 | 0.76 | 86 | 0.00 |
| Cough | 22 | 65.2 (58.6-71.2) | 1689/2515 | 0.80 | 85 | 0.00 |
| Dyspnea | 13 | 27.4 (19.6-35.2) | 477/2014 | 0.42 | 89 | 0.00 |
| Myalgia/fatigue | 17 | 34.7 (26.0-44.4) | 742/1938 | 0.60 | 89 | 0.00 |
| Sputum production | 12 | 17.2 (10.8-26.4) | 480/1862 | 0.01 | 89 | 0.00 |
| Sore throat | 7 | 14.5 (10.6-19.5) | 224/1577 | 0.88 | 66 | 0.00 |
| Headache | 12 | 11.1 (7.7-15.7) | 230/1864 | 0.30 | 74 | 0.00 |
| Diarrhea | 13 | 5.7 (3.8-8.6) | 104/2041 | 0.77 | 66 | 0.00 |
| Hemoptysis | 4 | 2.4 (0.8-6.7) | 20/1339 | 0.77 | 100 | 0.00 |
| Anorexia | 4 | 10.1 (1.0-57.2) | 82/1322 | 0.73 | 98 | 0.00 |
| Nausea/vomiting | 7 | 5.0 (2.3-10.7) | 65/1563 | 0.90 | 85 | 0.00 |
| Dizziness | 3 | 8.6 (2.5-26.0) | 16/205 | 0.90 | 65 | 0.00 |
| Chest tightness | 5 | 8.4 (2.5-26.0) | 24/256 | 0.24 | 78 | 0.00 |
| Rhinorrhea | 3 | 9.3 (2.2-31.0) | 28/232 | 0.17 | 88 | 0.00 |
| Chills | 2 | 14.3 (3.0-47.4) | 12/111 | NA | 86 | 0.00 |

### Laboratory findings

The most frequent abnormal laboratory findings in patients with COVID-19 were respectively, increased C-Reactive Protein (CRP) (72% CI 54.3-84.6), lymphopenia (50.1%, CI 38.0-62.4), high levels of Lactate Dehydrogenase (LDH) (41%,CI 22.8-62.0), increased serum aspartate aminotransferase (19.7%, CI 10.5-33.7) and thrombocytopenia (11.1%, CI 7.7-15.7) (Table 4). Among the confirmed COVID19 subjects, 14.0% (CI, 6.7-29.0) had viremia. Impaired hepatic function with ALT levels greater than 47.25 U/L was seen in 13.3% (CI 3.2-41.0) of COVID19 subjects. Acute cardiac injury with troponin levels greater than 28 pg/ml was seen in 12.4% of the patients. Acute kidney injury was found in 5.5% (CI 1.3-20.8). Shock was reported in 4.0% (CI 1.6-12.0) shock. 13.0% (CI 4.8-30.0) met the definition of acute respiratory distress syndrome (ARDS).

**Table 4.** Meta-analysis of laboratory findings.

|  | Number<br>of studies | Pooled<br>frequency<br>(95% CI) | n/N* | Publication<br>bias<br>(*p* value) | Heterogeneity test |  |
| --- | --- | --- | --- | --- | --- | --- |
|  |  |  |  |  | I<sup>2</sup> (%) | *p* value |
| Lymphopenia | 11 | 50.1 (38.0-62.4) | 1122/1853 | 0.08 | 93 | 0.00 |
| Lymphocytosis | 2 | 33.5 (2.4-90.2) | 55/93 | NA | 88 | 0.00 |
| Neutrophilia | 3 | 29.7 (19.3-42.7) | 60/191 | 0.51 | 58.7 | 0.08 |
| Leukopenia | 9 | 28.0 (20.0-37.4) | 544/1798 | 0.89 | 88 | 0.00 |
| Leukocytosis | 9 | 10.8 (5.8-19.1) | 165/1829 | 0.86 | 92 | 0.00 |
| Thrombocytopenia | 6 | 11.1 (7.7-15.7) | 343/1393 | 0.00 | 86 | 0.00 |
| Anemia | 2 | 43.5 (30.3-57.7) | 79/179 | NA | 72 | 0.00 |
| Decreased Albumin | 3 | 51.8 (2.0-98.0) | 105/191 | 0.99 | 96 | 0.00 |
| High CRP | 8 | 72.0 (54.3-84.6) | 918/1681 | 0.02 | 96 | 0.00 |
| High LDH | 6 | 41.0 (22.8-62.0) | 408/1393 | 0.32 | 94 | 0.00 |
| High ESR | 2 | 79.7 (66.6-88.5) | 143/179 | NA | 69 | 0.00 |
| High AST | 7 | 19.7 (10.5-33.7) | 267/1474 | 0.70 | 93 | 0.00 |
| High ALT | 4 | 14.6 (7.6-26.3) | 191/1290 | 0.99 | 84.8 | 0.00 |
| High Creatinine Kinase | 7 | 14.1 (8.3-23.0) | 142/1453 | 0.20 | 84 | 0.00 |
| High Bilirubin | 3 | 7.9 (2.9-19.0) | 95/1278 | 0.96 | 89 | 0.00 |
| High Creatinine | 5 | 3.3 (1.2-9.1) | 20/1294 | 0.13 | 74 | 0.00 |
| High Troponin I | 1 | 2.4 (0.3-15.0) | 1/41 | NA | 0.00 | 0.1 |

### Radiologic findings

Chest X-Ray (CXR) and chest CT-scan were the common imaging modalities used for the diagnosis of COVID19. The pooled sensitivity for CT-scan for COVID19 was 79.3%. The most common sites of the lung involvement based on chest CT-scan were right lower lobe (76.2, CI 57.8-82.5) followed by left lower lobe (71.8%, CI 57.8-82.5). Most of the patients (74.8%) had bilateral involvement. The most common pattern of parenchymal involvement was ground-glass opacity (66.0%, CI 51.1-78.0). The chest CT-scan was reported normal in 20.7% of the patients with confirmed RT-PCR results (Table 5).

**Table 5.** Meta-analysis of imaging findings.

| CT Scan | Patterns |  | Number of studies | Pooled frequency (95% CI) | n/N* | Publication bias (p value) | Heterogeneity test |  |
| --- | --- | --- | --- | --- | --- | --- | --- | --- |
|  |  |  |  |  |  |  | I<sup>2</sup> (%) | p value |
| Location of involvement | Number of affected lobe | Unaffected | 3 | 20.7 (15.1-27.6) | 33/161 | 0.18 | 0.0 | 0.57 |
|  |  | 1 lobe | 5 | 14.8 (7.4-24.0) | 52/318 | 0.22 | 73 | 0.00 |
|  |  | 2 lobes | 5 | 9.5 (6.5-12.8) | 30/318 | 0.32 | 0.0 | 0.50 |
|  |  | 3 lobes | 5 | 11.7 (7.9-14.6) | 36/318 | 0.64 | 0.0 | 0.50 |
|  |  | 4 lobes | 5 | 15.8 (10.3-20.7) | 49/318 | 0.90 | 40 | 0.15 |
|  |  | 5 lobes | 5 | 37.2 (32.0-42.3) | 118/318 | 0.50 | 30 | 0.22 |
|  | Affected lobe (s) | RUL | 4 | 56.8 (50.6-62.8) | 145/255 | 0.12 | 52 | 0.10 |
|  |  | RML | 4 | 48.6 (42.5-54.8) | 124/255 | 0.07 | 0.0 | 0.48 |
|  |  | RLL | 4 | 76.2 (65.5-84.4) | 193/255 | 0.14 | 64 | 0.03 |
|  |  | LUL | 4 | 56.0 (47.1-64.7) | 153/255 | 0.12 | 0.0 | 0.40 |
|  |  | LLL | 3 | 71.8 (57.8-82.5) | 167/234 | 0.30 | 76 | 0.01 |
|  | Laterality | Uni lateral | 3 | 28.8 (16.6-45.2) | 62/205 | 0.80 | 77 | 0.01 |
|  |  | Bi lateral | 3 | 70.6 (55.3-82.5) | 142/205 | 0.20 | 74 | 0.01 |
| Pattern of involvement | Pattern of involvement | No involvement | 4 | 17.2 (11.4-25.0) | 193/1080 | 0.42 | 63.0 | 0.04 |
|  |  | Both of GGO* & Consolidation | 2 | 39.0 (28.1-51.0) | 57/142 | NA | 25 | 0.24 |
|  |  | GGO without consolidation | 10 | 66.0 (51.1-78.0) | 846/1365 | 0.67 | 90 | 0.00 |
|  |  | Consolidation without GGO | 3 | 9.4 (3.3-23.6) | 26/274 | 0.21 | 82 | 0.00 |
|  | Laterality | Uni lateral | 7 | 21.8 (12.0-36.3) | 101/507 | 0.63 | 87 | 0.00 |
|  |  | Bi lateral | 8 | 74.8 (62.5-84.0) | 405/548 | 0.29 | 84 | 0.00 |

\*GGO: Ground Glass Opacities

### Outcomes

Hospitalization was required in 94.6% of patients with severe COVID-19. The pooled mortality rate of these patients was about 6.6% (CI 2.8-15.0) (Table 6, 7). Old age, male sex, presence of underlying diseases, higher level of D-dimer, lower level of fibrinogen and anti-thrombin, progressive radiographic deterioration in follow up CT-scans, developed ARDS and requirement of mechanical ventilation were reported factors associated with increased mortality rate. As shown in Table 8, men had significantly higher mortality in the hospital compared to women (OR 3.4; 95% CI 1.2-9.1, P = 0.01). Although ICU admission was higher in men, the difference was not significant. The mean duration between the time of hospitalization and death was 17.5 days with minimum and maximum periods of 14 and 21 days respectively. The effects and summaries calculated using a random-effects model weighted by the study population is shown in Figure 2.

**Table 6.** Meta-analysis of complications.

|  | Number of studies | Pooled frequency (95% CI) | n/N* | Publication bias (*p* value) | Heterogeneity test |  |
| --- | --- | --- | --- | --- | --- | --- |
|  |  |  |  |  | I<sup>2</sup> (%) | *p* value |
| RNAemia | 1 | 14.0 (6.7-29.0) | 6/41 | NA | 0.00 | 1.00 |
| ARDS | 9 | 13.0 (4.8-30.0) | 142/1794 | 0.67 | 96 | 0.00 |
| Acute cardiac injury | 4 | 12.4 (6.2-23.2) | 28/243 | 0.83 | 65 | 0.03 |
| Acute kidney injury | 6 | 5.5 (1.3-20.8) | 34/1441 | 0.58 | 93 | 0.00 |
| Liver failure | 3 | 13.3 (3.2-41.0) | 20/144 | 0.50 | 84 | 0.00 |
| Shock | 5 | 4.0 (1.6-12.0) | 32/1389 | 0.60 | 86 | 0.00 |
| Hospitalization | 10 | 94.6 (73.8-99.1) | 1561/1829 | 0.76 | 98 | 0.00 |

**Table 7.** Meta-analysis of outcomes.

|  | Number<br>of<br>studies | Pooled frequency<br>(95% CI) | n/N* | Publication<br>bias<br>(*p* value) | Heterogeneity test |  |
| --- | --- | --- | --- | --- | --- | --- |
|  |  |  |  |  | I<sup>2</sup> (%) | *p* value |
| Discharged | 14 | 52.7 (36.5-68.4) | 486/948 | 0.44 | 93 | 0.00 |
| Death | 11 | 6.6 (2.8-15.0) | 111/2026 | 0.50 | 93 | 0.00 |

**Table 8.** Mortality and ICU admission in men vs women in patients with COVID-19.

|  | Number<br>of studies | Pooled OR<br>(95% CI) | *p* value | Heterogeneity test |  |
| --- | --- | --- | --- | --- | --- |
|  |  |  |  | I<sup>2</sup> (%) | *p* value |
| Mortality in men vs women | 2 | 3.4 (1.2-9.1) | 0.01 | 0.00 | 0.6 |
| ICU admission in men vs women | 2 | 1.6 (0.7-3.2) | 0.1 | 0.00 | 0.5 |

**Figure 2.**
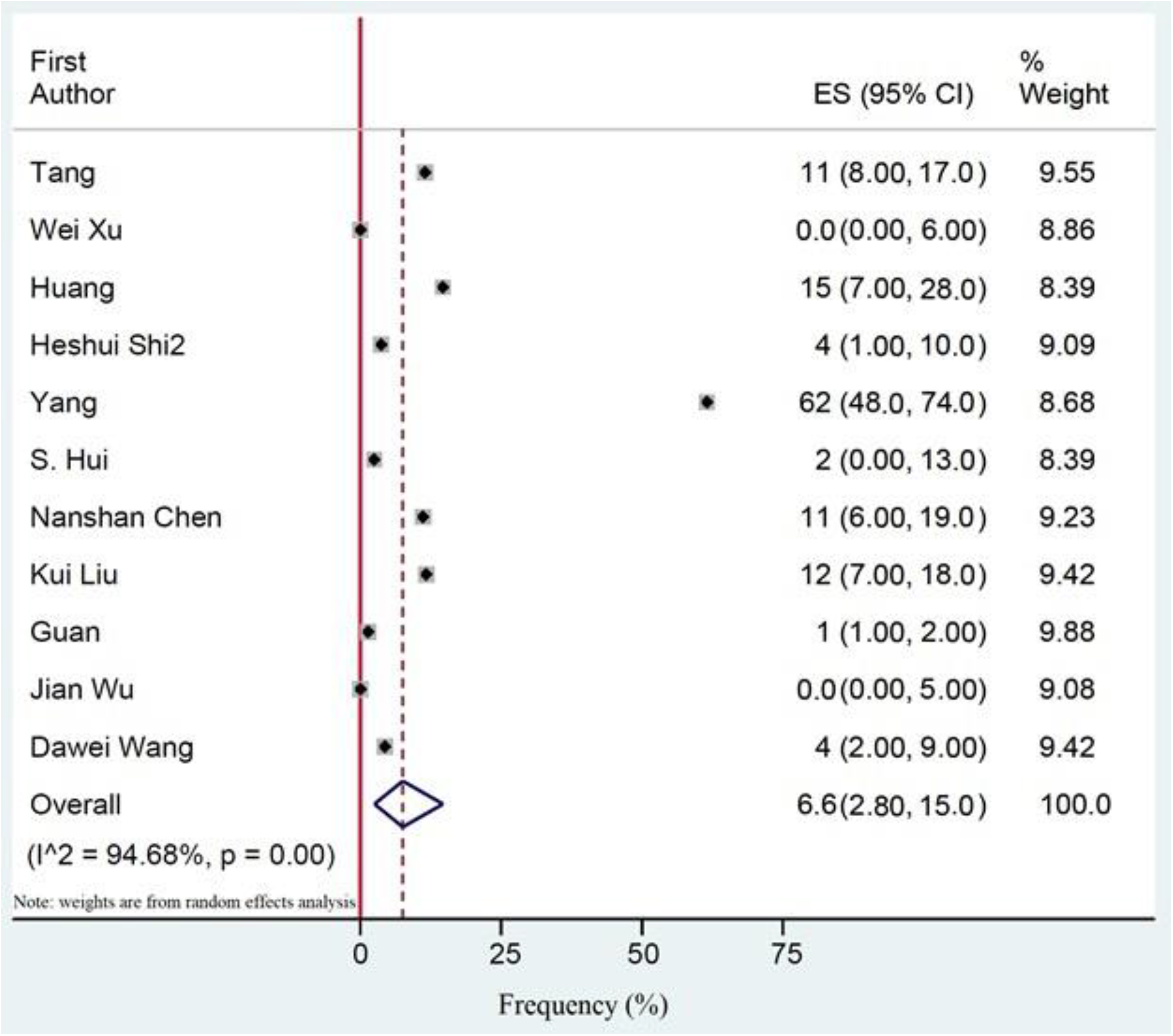
The pooled mortality rate of patients with COVID-19. Effects and summaries were calculated using a random-effects model weighted by study population.

## Discussion

We evaluated the signs and symptoms, diagnostic modalities, therapeutic measures and epidemiologic features of COVID-19 to have a better understanding of this pandemic caused by SARS-CoV-2. In terms of clinical manifestations, the most common signs and symptoms were fever and cough. Hypertension and cardiovascular diseases were the most common comorbidities among patients. Between the different abnormal laboratory findings, increased levels of CRP and lymphopenia were the most common findings. Chest X-Ray and chest CT-scan were the most common imaging modalities used for the diagnosis. The pooled sensitivity of CT-san for COVID19 was 79.3%. We found 20.7% of the patients with confirmed RT-PCR who had normal chest CT-Scan suggesting that a normal chest CT-scan cannot rule out the disease in patients who are highly suspicious for COVID-19. Several complications were seen due to COVID-19. Among these, acute hepatitis was the most common one occurring in 13.3% of cases, followed by cardiac injury with troponin levels greater than 28 pg/ml seen in 12.4%. The pooled mortality rate of these patients was 6.6%. We detected several factors contributing to a worse outcome including old age, male sex, presence of underlying diseases and some abnormal laboratory finding such as high level of D-Dimer. Although there was not any significant difference between male and female gender in ICU admissions, male gender showed a significantly higher in-hospital mortality rate.

The current study showed that fever (83.0%), cough (65.2%) and dyspnea (27.4%) were the most common signs and symptoms. In a study done by Zhang et al in Wuhan, fever was identified as the most common clinical finding present in 91.7% of the patients followed by cough in 75% of patients. Their study showed a higher gastrointestinal (GI) manifestation at presentation of the disease, representing 39.6% of the patients (10). Our study showed a lower prevalence of GI symptoms including diarrhea, which was present in 5.7% of patients and nausea/vomiting in 5%.

The most prevalent comorbidities in our study were hypertension (18.5 %, CI 12.7-24.4) followed by cardiovascular diseases (14.9 %, CI 6.0-23.8) and diabetes (10.8 %, CI 8.3-13.3). According to a systematic review for comorbidities by Yang et al, hypertension (17 ± 7%, CI 14-22%) and diabetes (8 ± 6%, CI 6-11%), followed by cardiovascular diseases (5 ± 4%, CI 4-7%) were the most common comorbid findings (11). The high prevalence of hypertension and other cardiovascular comorbidities have raised speculation regarding the role of angiotensin-converting enzyme Inhibitors (ACEI) in COVID-19. Angiotensin-converting enzyme 2 (ACE2) receptor has been identified as the receptor used by SARS-CoV-2 to infect human cells and previous studies have shown that the usage of ACEI results in the upregulation of ACE2 (12) (13). Theoretically, this increase in ACE2 levels could result in a greater risk of infection with the SARS-CoV-2 virus. Current evidence against the use of ACEI in patients with COVID-19 or those at risk of the disease is limited, and further studies are needed to analyze this possible association.

The most common laboratory abnormalities were elevated C-Reactive Protein (CRP) (72%), lymphopenia (50.1%) and elevated LDH (41%). Also, thrombocytopenia was seen in 11.1% of the subjects. Analyses by Huang et al and Lippi et al have suggested that lymphopenia and thrombocytopenia in COVID-19 patients are associated with poorer outcomes, respectively (14, 15). Interestingly, patients with SARS were reported to have a higher percentage of lymphopenia (68-90%) and thrombocytopenia (20-45%) compared to COVID-19 patients (16). A similar reduction in counts was also observed in patients with H1N1 influenza (17). Thrombocytopenia and lymphopenia have been previously shown to strongly indicate a higher risk of mortality in SARS and influenza (17, 18). Given the current lack of predictive biomarkers in COVID-19, lymphocyte and platelet count may be used as indicators of severe disease in the clinical setting.

Liver abnormality was the most common complication and was present among 13.3% of the subjects, although the data was reported only in 3 studies. However, a significant number of subjects had elevated AST (19.7%) and ALT (14.6%). Impaired liver function has been observed as collateral damage in some viral infections including SARS, possibly caused by direct damage to the hepatic tissue by the pathogen (19). While this could be the case with COVID19, an iatrogenic effect due to medications like lopinavir cannot be ruled out.

Another significant finding in our analysis was the incidence of cardiac injury in 12.4% of the patients, which is a common event seen in a multitude of viral illnesses(20). Gao et al observed that subjects with influenza (H7N9) and cardiac injury had an elevated risk of mortality (HR=2.06) (21). In a study by Ludwig et al for analysis of cardiac biomarkers in influenza patients, 24% of the subjects showed ACI in ≤30 days after influenza diagnosis and half of them were myocardial infarction (22). Although our analysis did not show increased mortality risk in patients with cardiac injury, these findings could indicate the potential need for identifying and optimizing cardiac risk factors in COVID-19 patients during the treatment period.

The mean duration between hospitalization and death was 17.5 days (range-14-21 days), compared to 17.4 days in SARS (23). The overall mortality rate in this study was 6.6%, which is more than twice than previously reported (24). Though comparable mortality was reported by Li et al (7%) and Qian et al (8.9%) in their meta-analyses, a study by Rodriguez et al showed a much higher death rate of 13.9% (25-27). On the other hand, a study from the Jiangsu province of China results showed a high cure rate equal to 96.67%. Although the main reason for very low mortality on this study remains unknown, measures including early recognition and centered-quarantine may be contributing factors(28).

Of note, the in-hospital mortality of males was significantly higher than females (OR 3.4; 95% CI 1.2-9.1, P = 0.01). A similar pattern of higher mortality in males has been reported in previous coronavirus outbreaks of SARS and MERS. Karlberg et al also reported that the gender-based difference in mortality was higher in younger males (0-44 years) (RR=2), compared to those of age group 45-74 (RR-1.45) (6). Similarly, the study by Alghamdi et al showed that the case fatality rate in males was twice that of females in MERS (52% VS 23%) (29). Although a gender-based difference in the immune response to infections has been suggested as a possible factor, other contributing factors including smoking history and severity of underlying comorbidities cannot be ruled out (30). This is especially of significance in China, where the prevalence of smoking among men (57.6%) is almost 10 times higher than in women (6.7%) (31). This difference in mortality opens the discussion for the need to treat COVID19 more aggressively in males, including the possibility of earlier intubation and mechanical ventilation in this population. Further investigations are also needed to understand this phenomenon.

This study has several limitations. Due to the rapidly emerging COVID-19 situation around the globe and the novelty of this coronavirus, there is still limited clinical data regarding diagnostic modalities and effective therapeutic measures. Most of the clinical findings were from observational studies. Future clinical trials and animal models are also required to have conclusive clinical information. We also need more studies outside of China for a comprehensive result that reflects COVID-19 globally. In the end, due to the lack of accurate reports of the new cases in different countries, the epidemiologic measures are also limited. As this pandemic is growing fast, future studies are needed for the evaluation of epidemiologic and clinical features of COVID-19.

## Conclusion

COVID-19 presents with a significant number of mortalities especially among males around the world. The pooled mortality rate in our study was 6.6%. The high rate of hospitalization and case fatality among hospitalized patients along with the lack of intensive care facilities necessitated the identification of factors associated with the severe disease and mortality. These factors included male gender, age, underlying diseases, higher level of D-Dimer, lower level of fibrinogen and anti-thrombin, progressive radiographic deterioration on follow up, developed ARDS and requirement of mechanical ventilation. It was inferred that higher comorbidities such as hypertension and cardiovascular diseases could be related to the pathogenesis of the virus through ACE II receptor. This association could open doors for future studies to evaluate the role of ACE inhibitor drugs in the high-risk group. There are still a lot of unknown features of COVID-19 for the broad scientific community to investigate in an effort to slow the progression and mortality of COVID-19 and finally defeat this pandemic.

## Data Availability

Authors can confirm that all relevant data are included in the article and/or its supplementary information files

## References

1. Gates B. Responding to Covid-19 — A Once-in-a-Century Pandemic? New England Journal of Medicine. 2020.

2. Coronavirus disease 2019 (COVID-19) situation summary: Center for Disease Control and Prevention 2020 [cited 2020 3/12/2020]. Available from: https://www.cdc.gov/coronavirus/2019-ncov/summary.html.

3. Peeri NC, Shrestha N, Rahman MS, Zaki R, Tan Z, Bibi S, et al. The SARS, MERS and novel coronavirus (COVID-19) epidemics, the newest and biggest global health threats: what lessons have we learned? International Journal of Epidemiology. 2020.

4. Lukelsu. Coronavirus Dashboard (GEOG 4046 example) 2020 [Available from: https://www.arcgis.com/apps/opsdashboard/index.html#/90b89509931e4c309e83d7f51e101a08.

5. Wu C, Chen X, Cai Y, Xia J, Zhou X, Xu S, et al. Risk Factors Associated With Acute Respiratory Distress Syndrome and Death in Patients With Coronavirus Disease 2019 Pneumonia in Wuhan, China. JAMA Intern Med. 2020.

6. Karlberg J, Chong DS, Lai WY. Do men have a higher case fatality rate of severe acute respiratory syndrome than women do? Am J Epidemiol. 2004;159(3):229–31.

7. Alghamdi IG, Hussain, II, Almalki SS, Alghamdi MS, Alghamdi MM, El-Sheemy MA. The pattern of Middle East respiratory syndrome coronavirus in Saudi Arabia: a descriptive epidemiological analysis of data from the Saudi Ministry of Health. Int J Gen Med. 2014;7:417–23.

8. Moher D, Liberati A, Tetzlaff J, Altman DG. Preferred reporting items for systematic reviews and meta-analyses: the PRISMA statement. Annals of internal medicine. 2009;151(4):264–9.

9. Munn Z, Moola S, Lisy K, Riitano D. The Joanna Briggs institute reviewers’ manual 2014. The systematic review of prevalence and incidence data Adelaide: The Joanna Briggs Institute. 2014.

10. Zhang Jj, Dong X, Cao YY, Yuan Yd, Yang Yb, Yan Yq, et al. Clinical characteristics of 140 patients infected by SARS-CoV-2 in Wuhan, China. Allergy. 2020.

11. Yang J, Zheng Y, Gou X, Pu K, Chen Z, Guo Q, et al. Prevalence of comorbidities in the novel Wuhan coronavirus (COVID-19) infection: a systematic review and meta-analysis. Int J Infect Dis. 2020.

12. Zhang H, Penninger JM, Li Y, Zhong N, Slutsky AS. Angiotensin-converting enzyme 2 (ACE2) as a SARS-CoV-2 receptor: molecular mechanisms and potential therapeutic target. Intensive Care Medicine. 2020.

13. Tikellis C, Thomas MC. Angiotensin-Converting Enzyme 2 (ACE2) Is a Key Modulator of the Renin Angiotensin System in Health and Disease. Int J Pept. 2012;2012:256294.

14. Huang C, Wang Y, Li X, Ren L, Zhao J, Hu Y, et al. Clinical features of patients infected with 2019 novel coronavirus in Wuhan, China. Lancet. 2020;395(10223):497–506.

15. Lippi G, Plebani M, Michael Henry B. Thrombocytopenia is associated with severe coronavirus disease 2019 (COVID-19) infections: A meta-analysis. Clin Chim Acta. 2020.

16. Yang M, Hon KL, Li K, Fok TF, Li CK. The effect of SARS coronavirus on blood system: its clinical findings and the pathophysiologic hypothesis. Zhongguo Shi Yan Xue Ye Xue Za Zhi. 2003;11(3):217–21.

17. Lopez-Delgado JC, Rovira A, Esteve F, Rico N, Manez Mendiluce R, Ballus Noguera J, et al. Thrombocytopenia as a mortality risk factor in acute respiratory failure in H1N1 influenza. Swiss Med Wkly. 2013;143:w13788.

18. Bellelli V, d’Ettorre G, Celani L, Borrazzo C, Ceccarelli G, Venditti M. Clinical significance of lymphocytopenia in patients hospitalized with pneumonia caused by influenza virus. Critical Care. 2019;23(1):330.

19. Peiris JSM, Lai ST, Poon LLM, Guan Y, Yam LYC, Lim W, et al. Coronavirus as a possible cause of severe acute respiratory syndrome. The Lancet. 2003;361(9366):1319–25.

20. Greaves K, Oxford JS, Price CP, Clarke GH, Crake T. The Prevalence of Myocarditis and Skeletal Muscle Injury During Acute Viral Infection in Adults: Measurement of Cardiac Troponins I and T in 152 Patients With Acute Influenza Infection. Archives of Internal Medicine. 2003;163(2):165–8.

21. Gao C, Wang Y, Gu X, Shen X, Zhou D, Zhou S, et al. Association Between Cardiac Injury and Mortality in Hospitalized Patients Infected With Avian Influenza A (H7N9) Virus. Read Online: Critical Care Medicine | Society of Critical Care Medicine. 2020;48(4):451–8.

22. Ludwig A, Lucero-Obusan C, Schirmer P, Winston C, Holodniy M. Acute cardiac injury events </=30 days after laboratory-confirmed influenza virus infection among U.S. veterans, 2010-2012. BMC Cardiovasc Disord. 2015;15:109.

23. Feng D, Jia N, Fang L-Q, Richardus JH, Han X-N, Cao W-C, et al. Duration of symptom onset to hospital admission and admission to discharge or death in SARS in mainland China: a descriptive study. Tropical Medicine & International Health. 2009;14(1):28–35.

24. Guan W-j, Ni Z-y, Hu Y, Liang W-h, Ou C-q, He J-x, et al. Clinical Characteristics of Coronavirus Disease 2019 in China. New England Journal of Medicine. 2020.

25. Qian K, Deng Y, Tai Y, Peng J, Peng H, Jiang L. Clinical Characteristics of 2019 Novel Infected Coronavirus Pneumonia:A Systemic Review and Meta-analysis. medRxiv. 2020:2020.02.14.20021535.

26. Li L-q, Huang T, Wang Y-q, Wang Z-p, Liang Y, Huang T-b, et al. 2019 novel coronavirus patients’ clinical characteristics, discharge rate and fatality rate of meta-analysis. Journal of Medical Virology. n/a(n/a).

27. Rodriguez-Morales AJ, Cardona-Ospina JA, Gutiérrez-Ocampo E, Villamizar-Peña R, Holguin-Rivera Y, Escalera-Antezana JP, et al. Clinical, laboratory and imaging features of COVID-19: A systematic review and meta-analysis. Travel Medicine and Infectious Disease. 2020:101623.

28. Sun Q, Qiu H, Huang M, Yang Y. Lower mortality of COVID-19 by early recognition and intervention: experience from Jiangsu Province. Annals of Intensive Care. 2020;10(1):33.

29. Alghamdi IG, Hussain II, Almalki SS, Alghamdi MS, Alghamdi MM, El-Sheemy MA. The pattern of Middle East respiratory syndrome coronavirus in Saudi Arabia: a descriptive epidemiological analysis of data from the Saudi Ministry of Health. Int J Gen Med. 2014;7:417–23.

30. Klein SL. The effects of hormones on sex differences in infection: from genes to behavior. Neuroscience & Biobehavioral Reviews. 2000;24(6):627–38.

31. Yang T, Barnett R, Jiang S, Yu L, Xian H, Ying J, et al. Gender balance and its impact on male and female smoking rates in Chinese cities. Social Science & Medicine. 2016;154:9–17.

32. Hui DS, I Azhar E, Madani TA, Ntoumi F, Kock R, Dar O, et al. The continuing 2019-nCoV epidemic threat of novel coronaviruses to global health—The latest 2019 novel coronavirus outbreak in Wuhan, China. International Journal of Infectious Diseases. 2020;91:264–6.

33. Xia J, Tong J, Liu M, Shen Y, Guo D. Evaluation of coronavirus in tears and conjunctival secretions of patients with SARS-CoV-2 infection. Journal of Medical Virology. 2020.

34. Xu X-W, Wu X-X, Jiang X-G, Xu K-J, Ying L-J, Ma C-L, et al. Clinical findings in a group of patients infected with the 2019 novel coronavirus (SARS-Cov-2) outside of Wuhan, China: retrospective case series. Bmj. 2020;368.

35. Zhang W, Du R-H, Li B, Zheng X-S, Yang X-L, Hu B, et al. Molecular and serological investigation of 2019-nCoV infected patients: implication of multiple shedding routes. Emerging microbes & infections. 2020;9(1):386–9.

36. To KK-W, Tsang OTY, Chik-Yan Yip C, Chan K-H, Wu T-C, Chan J, et al. Consistent detection of 2019 novel coronavirus in saliva. Clinical Infectious Diseases. 2020.

37. Zou L, Ruan F, Huang M, Liang L, Huang H, Hong Z, et al. SARS-CoV-2 viral load in upper respiratory specimens of infected patients. New England Journal of Medicine. 2020.

38. Hoehl S, Berger A, Kortenbusch M, Cinatl J, Bojkova D, Rabenau H, et al. Evidence of SARS-CoV-2 infection in returning travelers from Wuhan, China. New England Journal of Medicine. 2020.

39. Pan Y, Zhang D, Yang P, Poon LL, Wang Q. Viral load of SARS-CoV-2 in clinical samples. The Lancet Infectious Diseases. 2020.

40. Tang N, Li D, Wang X, Sun Z. Abnormal Coagulation parameters are associated with poor prognosis in patients with novel coronavirus pneumonia. Journal of Thrombosis and Haemostasis. 2020.

41. Chung M, Bernheim A, Mei X, Zhang N, Huang M, Zeng X, et al. CT imaging features of 2019 novel coronavirus (2019-nCoV). Radiology. 2020:200230.

42. Fang Y, Zhang H, Xie J, Lin M, Ying L, Pang P, et al. Sensitivity of chest CT for COVID-19: comparison to RT-PCR. Radiology. 2020:200432.

43. Guan W-j, Ni Z-y, Hu Y, Liang W-h, Ou C-q, He J-x, et al. Clinical characteristics of coronavirus disease 2019 in China. New England Journal of Medicine. 2020.

44. Huang C, Wang Y, Li X, Ren L, Zhao J, Hu Y, et al. Clinical features of patients infected with 2019 novel coronavirus in Wuhan, China. The Lancet. 2020;395(10223):497–506.

45. Kui L, Fang Y-Y, Deng Y, Liu W, Wang M-F, Ma J-P, et al. Clinical characteristics of novel coronavirus cases in tertiary hospitals in Hubei Province. Chinese medical journal. 2020.

46. Li Q, Guan X, Wu P, Wang X, Zhou L, Tong Y, et al. Early transmission dynamics in Wuhan, China, of novel coronavirus–infected pneumonia. New England Journal of Medicine. 2020.

47. Liu Y, Yang Y, Zhang C, Huang F, Wang F, Yuan J, et al. Clinical and biochemical indexes from 2019-nCoV infected patients linked to viral loads and lung injury. Science China Life Sciences. 2020:1–11.

48. Wang D, Hu B, Hu C, Zhu F, Liu X, Zhang J, et al. Clinical characteristics of 138 hospitalized patients with 2019 novel coronavirus–infected pneumonia in Wuhan, China. Jama. 2020.

49. Wu J, Liu J, Zhao X, Liu C, Wang W, Wang D, et al. Clinical Characteristics of Imported Cases of COVID-19 in Jiangsu Province: A Multicenter Descriptive Study. Clinical Infectious Diseases. 2020.

50. Ai T, Yang Z, Hou H, Zhan C, Chen C, Lv W, et al. Correlation of chest CT and RT-PCR testing in coronavirus disease 2019 (COVID-19) in China: A report of 1014 cases. Radiology. 2020:200642.

51. Pan F, Ye T, Sun P, Gui S, Liang B, Li L, et al. Time course of lung changes on chest CT during recovery from 2019 novel coronavirus (COVID-19) pneumonia. Radiology. 2020:200370.

52. Zhu N, Zhang D, Wang W, Li X, Yang B, Song J, et al. A Novel Coronavirus from Patients with Pneumonia in China, 2019. New England Journal of Medicine. 2020;382(8):727–33.

53. Yang X, Yu Y, Xu J, Shu H, Liu H, Wu Y, et al. Clinical course and outcomes of critically ill patients with SARS-CoV-2 pneumonia in Wuhan, China: a single-centered, retrospective, observational study. The Lancet Respiratory Medicine. 2020.

54. Bajema KL, Oster AM, McGovern OL, Lindstrom S, Stenger MR, Anderson TC, et al. Persons Evaluated for 2019 Novel Coronavirus—United States, January 2020. Morbidity and Mortality Weekly Report. 2020;69(6):166.

55. Bernheim A, Mei X, Huang M, Yang Y, Fayad ZA, Zhang N, et al. Chest CT findings in coronavirus disease-19 (COVID-19): Relationship to duration of infection. Radiology. 2020:200463.

56. Chen N, Zhou M, Dong X, Qu J, Gong F, Han Y, et al. Epidemiological and clinical characteristics of 99 cases of 2019 novel coronavirus pneumonia in Wuhan, China: a descriptive study. The Lancet. 2020;395(10223):507–13.

57. Pan Y, Guan H, Zhou S, Wang Y, Li Q, Zhu T, et al. Initial CT findings and temporal changes in patients with the novel coronavirus pneumonia (2019-nCoV): a study of 63 patients in Wuhan, China. European radiology. 2020:1–4.

58. Xu Y-H, Dong J-H, An W-m, Lv X-Y, Yin X-P, Zhang J-Z, et al. Clinical and computed tomographic imaging features of Novel Coronavirus Pneumonia caused by SARS-CoV-2. Journal of Infection. 2020.

59. Xu X, Yu C, Qu J, Zhang L, Jiang S, Huang D, et al. Imaging and clinical features of patients with 2019 novel coronavirus SARS-CoV-2. European Journal of Nuclear Medicine and Molecular Imaging. 2020:1–6.

60. Chang D, Lin M, Wei L, Xie L, Zhu G, Cruz CSD, et al. Epidemiologic and clinical characteristics of novel coronavirus infections involving 13 patients outside Wuhan, China. Jama. 2020.

61. Chen W, Lan Y, Yuan X, Deng X, Li Y, Cai X, et al. Detectable 2019-nCoV viral RNA in blood is a strong indicator for the further clinical severity. Emerging Microbes & Infections. 2020;9(1):469–73.

62. Kwok KO, Wong V, Wei VWI, Wong SYS, Tang JW-T. Novel coronavirus (2019-nCoV) cases in Hong Kong and implications for further spread. Journal of Infection. 2020.

